# Association of Salivary Biomarker Concentration and Activity Between Caries-free and Caries-affected Children: An Umbrella Review

**DOI:** 10.1101/2025.05.28.25328534

**Authors:** David Okuji, Danial Ahmed, Yulia Eve, Nicole Scott, Amir Yavari

## Abstract

**Objectives:** This umbrella review evaluates systematic reviews and meta-analyses for salivary biomarker concentration and activity in caries-affected versus caries-free children.

**Methods:** A comprehensive literature search identified relevant reviews, which were systematically selected using PRISMA guidelines, assessed qualitatively with AMSTAR 2, and analyzed quantitatively using RevMan software. Certainty of evidence was evaluated via the GRADE assessment tool.

**Results:** Of 609 identified articles, three reviews were included for quantitative analysis. AMSTAR 2 assessments rated three reviews as high quality and one as low quality. Meta-analysis findings showed that for salivary secretory immunoglobulin-A concentration with a mean of 65.54, with a 2.24 higher concentration (0.59 to 3.89 higher) in caries-affected children; carbonic anhydrase-VI concentration with a mean of 2.18, with a 0.92 lower concentration (2.21 lower to 0.38 higher) in caries-affected children; and carbonic anhydrase-VI activity with a mean of 3698.30, with a 2.89 higher activity level (1.24 to 4.54 higher) in caries-affected children. Heterogeneity was low for carbonic anhydrase, high for Salivary secretory immunoglobulin-A, and publication bias risk was low. The GRADE assessment indicated moderate confidence in evidence suggesting slight differences in Salivary secretory immunoglobulin-A and carbonic anhydrase-VI levels in caries-affected children.

**Conclusions:** Caries-affected children under age nine exhibit higher salivary secretory immunoglobulin-A concentration and carbonic anhydrase-VI activity but lower carbonic anhydrase-VI concentration. Current evidence suggests that screening with these three salivary biomarker tests are likely to benefit and unlikely to harm children. Widespread clinical application remains limited until U.S. commercial laboratories provide standardized saliva-based testing for salivary secretory immunoglobulin-A and carbonic anhydrase-VI.

## INTRODUCTION

### Background

Childhood dental caries is a prevalent, multifactorial disease affecting a significant proportion of children worldwide, including those under nine years old.^1^ Early childhood caries (**ECC**) poses serious risks, including pain, infection, and extensive restorative treatment, often requiring general anesthesia in young children.^2^ Current diagnostic methods, such as visual and radiographic examinations, lack sensitivity for early-stage detection before cavitation occurs, underscoring the need for improved diagnostic approaches.^3^

Saliva plays a fundamental role in oral health, contributing to buffering capacity, antimicrobial activity, and enamel remineralization.^4^ Because teeth are continuously bathed in saliva, its composition influences caries development. Cariogenic bacteria in the biofilm disrupt the balance of microbial communities, leading to mineral loss from dental hard tissues.^5^ Therefore, understanding salivary constituents provides valuable insight into caries progression.

Salivary biomarkers, which are measurable substances within saliva, offer a promising, non-invasive approach for detecting diseases, including dental caries.^6^ Biochemical and microbiological alterations in saliva may precede visible signs of ECC, making it an ideal medium for early-stage disease detection.^7^ Compared to traditional methods, salivary diagnostics offers advantages such as ease of collection, patient comfort, reduced infection risk, cost-effectiveness, and repeated sampling for long-term monitoring.^8^ Recent advancements in analytical technologies, including proteomics, genomics, metabolomics, microfluidics, and biosensors, have enhanced the identification and quantification of salivary biomarkers with high sensitivity and specificity.^9^ These innovations support the feasibility of point-of-care devices for ECC screening in dental offices, clinics, and even home settings, improving access to early diagnostics, particularly in underserved communities.^7^

### Rationale for Salivary Biomarker Testing for Early ECC Identification

Dental caries results from interactions among the host (tooth and saliva), cariogenic bacteria, and dietary carbohydrates.^2^ Bacterial fermentation of sugars generates organic acids, lowering pH and triggering enamel demineralization.^2^ Identifying salivary biomarkers that reflect these underlying biological processes in their early stages is crucial for preventive intervention.

Saliva provides protection through buffering acids, promoting enamel remineralization, clearing food debris, and exerting antimicrobial effects.^9^ Alterations in salivary flow rate, pH, buffering capacity, or key biomolecules may compromise this protective balance, increasing caries risk.^9^ Research has identified microbial species, proteins, enzymes, immunoglobulins, inflammatory mediators, and metabolic products as potential biomarkers for ECC detection.^9^

One key advantage of salivary biomarker testing is its ability to detect ECC in its earliest, pre-cavitated stages before visible signs appear.^9^ This early detection window allows timely preventive interventions to halt disease progression and reduce treatment burdens.^10^ Identifying children at elevated risk through salivary biomarkers enables personalized preventive strategies, including tailored oral hygiene guidance, dietary modifications, application of fluoride containing agents for enamel remineralization, and other minimally invasive care treatments.^2^ Risk-based prevention, guided by objective biomarker assessments, enhances specific and personalized preventive treatments compared to generalized interventions.^2^

Additionally, salivary biomarkers improve the accuracy of ECC risk assessment models by offering objective measures of susceptibility.^11^ Longitudinal studies tracking biomarker levels over time could further refine predictive capabilities.^11^ Given its non-invasive nature, saliva collection is ideal for young children, reducing anxiety and discomfort compared to traditional dental exams.^8^ Greater acceptability among children and caregivers supports improved participation in screening programs and facilitates routine monitoring.^6^

Finally, integrating salivary biomarker testing into early ECC detection frameworks could lead to more cost-effective disease management.^8^ By preventing lesion progression and minimizing the need for extensive restorative treatment, hospitalization, and emergency care, early intervention strategies could yield substantial financial savings for individuals, families, and healthcare systems.^2^

### Objective

The purpose of this study was to conduct an umbrella systematic review and determine if there are associations of salivary biomarker concentrations and activity between caries-affected (**CA**) and caries-free (**CF**) children under age nine years old. The rationale is to provide guidance to dentists with non-invasive screening tests for early identification for children who are at high risk for dental caries. The main research question for this umbrella review is: “Are there differences in specific biomarker concentration and/or activity between caries-free and caries-affected children?” The null hypothesis is there are no differences in specific salivary biomarker concentration and/or activity between CA and CF children.

## METHODS

### Research Protocol and Registration

This umbrella review was registered on 22^nd^ April 2024 and was assigned the identification number CRD42024538334 in the PROSPERO^12^ international prospective register of systematic reviews hosted by the National Institute for Health Research, Center for Reviews and Dissemination, University of York, UK.

#### Reporting format

This review was adapted and reported according to the Preferred Reporting Items for Systematic Reviews and Meta-Analysis (**PRISMA**)^13^ and the Cochrane Handbook for Systematic Reviews of Interventions^14^ throughout the process of this umbrella review.

### Eligibility criteria

#### Population, Intervention, Comparator, Outcome (*PICO)*

The population included healthy children who were less than nine years old. The indicator was caries-affected teeth, and the comparator was caries-free teeth, which were defined by either the International Caries Detection and Assessment System (ICDAS)^15^ or dmfs/DMFS and dmft/DMFT Index^16^ methods. The outcomes were mean average and standard deviation for concentration and/or activity of specific salivary biomarkers.

#### Criteria for selecting reviews for inclusion

The reviewed studies were designed as a systematic review, with meta-analysis of cross sectional or observational studies, which analyzed salivary protein abundance (concentration, activity or levels of total proteins, enzymes, immunoglobulins, antioxidants and antimicrobial peptides (**AMP**s) in dental plaque, saliva or acquired salivary film in children less than nine years old.

#### Exclusion Criteria

Reviewed studies not designed as a systematic review, with meta-analyses of cross-sectional, were excluded from this review. Articles which were classified as case reports, comments, laboratory studies, letters, and narrative reviews were excluded.

### Literature Search

#### Search Strategy, information sources, and selection process

An initial literature search was conducted on 3^rd^ September 2023 in all relevant publications in the PubMed, Embase, Cumulative Index to Nursing & Allied Health (CINAHL), Scopus, Web of Science, Cochrane Database of Systematic Reviews, Dentistry & Oral Sciences Source (**DOSS**) and, Health and Psychosocial Instruments (**HaPI**) databases, as well searching the grey literature. Hand searching and checking reference lists were also used to identify additional relevant records. The search strategy was composed of the following keywords and Boolean operators “systematic review,” “review,” or “meta-analysis,” using the keywords “children,” “salivary proteins,” “caries free,” and “caries affected” and applied filters for “meta-analysis,” “review,” “systematic review.” An updated search was conducted on 14^th^ April 2025, to identify additional studies.

#### Study selection

For the study selection, two selected authors independently screened the titles and abstracts using an Excel spreadsheet tool (Microsoft Corporation, Redmond, WA) to eliminate duplicates and read the full text of all papers to identify relevant systematic reviews, with meta-analyses. Discrepancies with selected reviews were resolved through discussion and mutual agreement by the two researchers.

The results of the study selection outcomes were populated into a Preferred Reporting Items for Systematic reviews and Meta-Analyses (PRISMA) flow diagram.^13^

### Data Collection and Analysis

#### Synthesis Methods

The methods utilized to synthesize the data from the reviewed articles included 1) Excel spreadsheet (Microsoft Corporation, Redmond, WA) to compile the characteristics of the included studies, 2) corrected covered area (**CCA**) citation matrix analysis to measure the degree of overlap, 3) “A MeaSurement Tool to Assess systematic Reviews” (**AMSTAR**) 2 to conduct the quality appraisal, risk of bias, heterogeneity of the data, 4) RevMan 5.4 software application *(The Cochrane Collaboration Review Manager 5, 2020, Copenhagen, Denmark)* to meta-analyze the effect size of variables, heterogeneity of the data, publication bias, and 5) Grading of Recommendations, Assessment, Development and Evaluation (**GRADE**) to assess the certainty of evidence (**COE**).

#### Data collection process and data items

Qualitative data extraction for the characteristics of the included systematic review and meta-analysis studies was completed by two of this study’s authors who independently focused on collecting details on the diagnostic tests, author names, publication year, journal name, research question, search strategies, study design, outcomes, assessment tools, and conclusions for each study marked for inclusion in this investigation. Disagreements were resolved through discussion and mutual agreement by the two researchers.

Quantitative data extraction was completed by the same two independent reviewers. For the meta-analysis summary for this part of the present umbrella review, the author names, year of publication, number and study design of the primary studies, point estimates with confidence intervals, heterogeneity statistics and quality appraisal, with RevMan 5.4 software (The Cochrane Collaboration Review Manager 5, 2020, Copenhagen, Denmark).

#### Analysis of degree of overlap

A citation matrix was generated to calculate the CCA. This analysis classifies the degree of overlap as “slight” (zero to five percent), “moderate” (six to ten percent), “high” (11 to 15 percent) or “very high” overlap (greater than 15 percent).^17^ The formula used is CCA = (N-r) / (rc-r) where “N” is number of included primary studies, “r” equals the number of index publications, and “c” includes the number of reviews.

#### Quality appraisal

The quality appraisal was conducted with the AMSTAR 2 tool, which enables the assessment of systematic reviews of randomized and non-randomized studies of healthcare interventions. The tool contains 16 domain items to assess the quality of included systematic reviews.^18^ The domain questions are designed so that a “yes” answer denotes a positive result and when information needed was present in the study. The “no” answer denotes when no information is available to rate. The “partial yes” answer denotes when partial information is provided in the article. The tool provides an overall rating based on weaknesses in critical domains.^18^

#### Risk of bias assessment

The risk of bias (**RoB**) assessment is included in two of the domain items of the AMSTAR 2 tool.^18^ The RoB from individual primary studies and from the interpretation of the results of the review are, respectively, included in domain “item 9” and “item 13” shown in Figure 2. “Item 15” in Figure 2 specifically assessed the risk of publication bias. Separately with the RevMan 5.4 software, publication bias was separately analyzed with funnel-plot analytic methodology, which is based on evidence for the effect of small-studies.

#### Estimation of common effect size

The primary aim of this umbrella review was to facilitate a straightforward comparison of the effects across various factors examined by utilizing a consistent measure of effect size. Since systematic reviews and meta-analyses incorporate different measures based on the design and analytical methods of the included studies, it was important to establish a common effect size to enable an overall comparison. Conversion of all effect size into mean average and mean difference was planned, using the RevMan 5.4 software.

#### Heterogeneity of data

The heterogeneity of the data was analyzed using the I² statistic which is quantified with the RevMan 5.4 software. In meta-analysis, the I² statistic quantifies the proportion of variability in a meta-analysis due to differences between included trials, rather than random sampling error. It ranges from 0% to 100%, with higher values indicating greater heterogeneity. As suggested guidelines, I² values can be categorized as low (25%), moderate (50%), and high (75%) heterogeneity.^19^

#### Certainty of evidence assessment

The certainty of evidence (**COE**) for quantitative outcome data utilized the Grading of Recommendations, Assessment, Development and Evaluation (**GRADE**)^20^ methodology to assess and report the confidence the investigators have in the effect estimate for each pre-defined, clinically important outcome of interest in the umbrella review. The investigators extracted and reported the GRADE assessments presented in the included systematic reviews and meta-analyses.

## RESULTS

### Description of included reviews

#### Literature search results and study selection

The search resulted in a total of 609 references, 493 sources from databases and registers, and 116 from the grey literature, that underwent screening.

All studies from the grey literature were excluded as 60 were duplicates and 56 did not meet the inclusion criteria. Of the 493 database- and register-sourced references, 150 were eliminated as duplicates. The 343 articles obtained were screened via their titles and abstracts, with 56 excluded as they did not conform with study objective and 3 were not retrieved. The remaining 284 articles were subjected to full text screening based on predefined eligibility criteria. Two hundred and sixty that did not meet the study inclusion criteria and were excluded from the study. The remaining 24 were further screened, and 21 studies were excluded because they did not meet the inclusion criteria.

After the two screening stages, a total of three systematic reviews with meta-analyses (Al-Mahdi *et al*.,^21^ da Silveira *et al*.,^22^ and Ravikumar *et al*.^23^) were included in this umbrella review. The details of the results of each search and screening stage can be found in PRISMA flow diagram (Figure 1).

**Figure 1.**
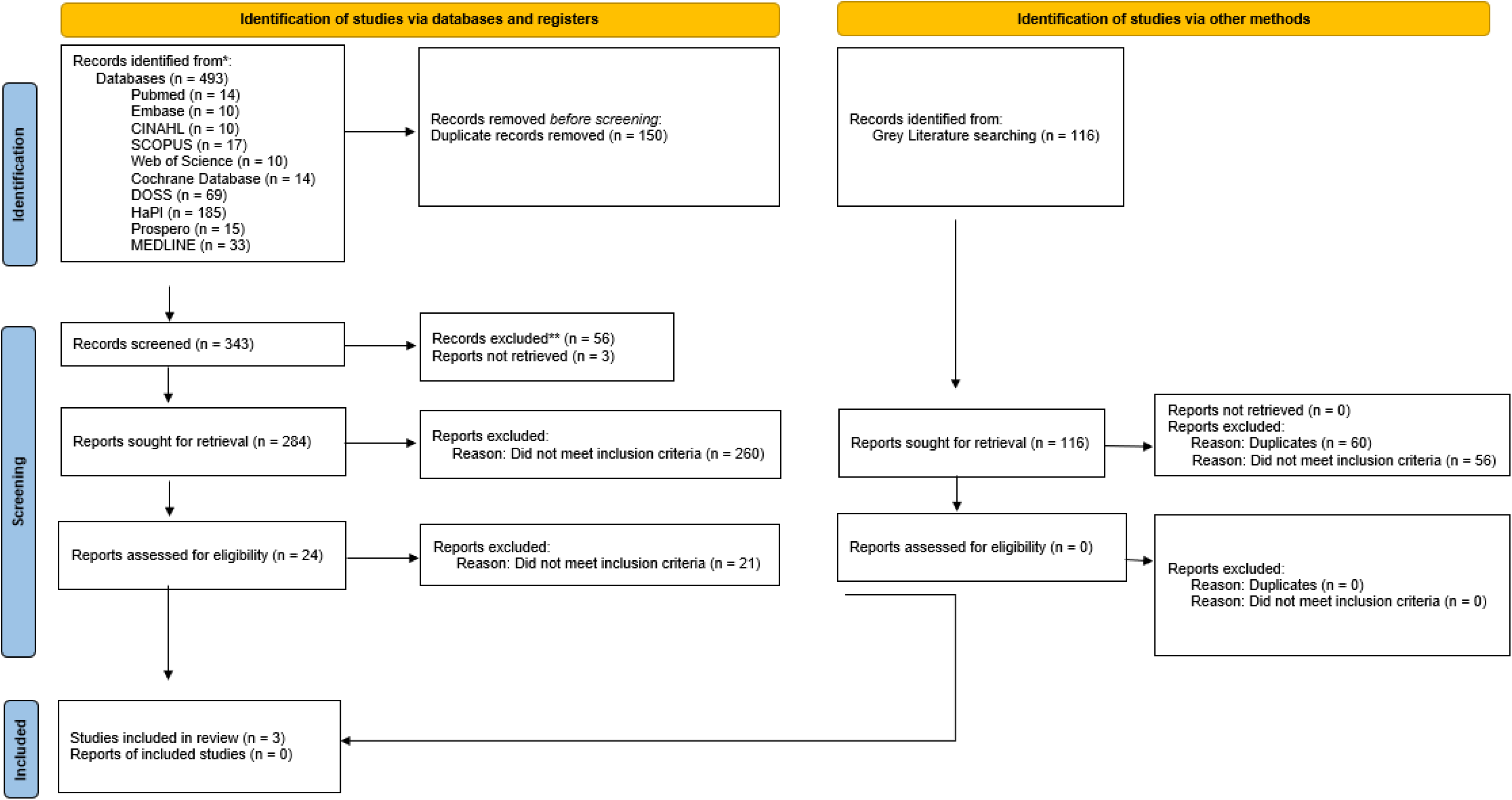
Preferred Reporting Items for Systematic reviews and Meta-Analyses (PRISMA) flow diagram of identification and selection of included reviews.

#### Study characteristics

As shown in Table 1, one review was conducted in Norway, Australia and Yemen,^21^ one in Brazil,^22^ and one in India.^23^ The number of databases searched by the included reviews ranged from five to nine, with the search period end-year ranging from 2021 to 2023.

**Table 1.** Characteristics of Included Articles.

| Authors, year, journal (country) | Search strategy: Databases | Search strategy: Search period start date | Search strategy: Search period end date | Search strategy: Language restriction | Study design of the included studies | Study design of the included studies: Number of studies | Research question | Study Population | Indicator/ Intervention | Comparator/ Control | Conclusions | Assessment tools | Meta-analysis performed |
| --- | --- | --- | --- | --- | --- | --- | --- | --- | --- | --- | --- | --- | --- |
| Al-Mahdi *et al.*, 2023, Clinical and Experimental Dental Research (Norway, Australia, Yemen) | 1. China National Knowledge Infrastructure (CNKI)<br>2. Cochrane Library<br>3. PubMed<br>4. Scopus<br>5. Web of Science | Not reported | 8/31/2022 | English or Chinese | Cross-sectional studies, Observational studies | Seven (7) included studies for meta-analysis, with all designed as cross-sectional studies | Is there any link between the concentration and/or activity of carbonic anhydrase-VI (CA-VI) and the susceptibility to dental caries? | For the seven (7) included meta-analysis studies, the study population included healthy children less than 9 years of age. | Children: Healthy with dental caries | Children: Healthy caries-free individuals | Activity and/or concentration of salivary or plaque carbonic anhydrase-VI (CA-VI) reported as the mean and standard deviation.<br>Carbonic anhydrase-VI (CA-VI) has lower concentrations and higher activity in patients with dental caries than in caries-free individuals. | Newcastle–Ottawa Scale (NOS) | Yes |
| da Silveira *et al.*, 2023, Frontiers in Oral Health (Brazil) | 1. Embase<br>2. International Scientific Index (ISI)<br>3. MEDLINE<br>4. Web of Science<br>5. Gray literature was not searched | Not reported | 2/28/2023 | English, German, or in Latin-based languages | Observational studies | All fifteen (15) included studies for meta-analysis were designed as observational studies | Does the oral cavity of caries-affected (CA) individuals has a different protein abundance compared with the oral cavity of caries-free (CF) ones? | Children: Healthy children up to age 9 years old | Children: Caries-affected | Children: Healthy caries-free individuals | Protein abundance (concentration, activity or levels of total proteins, enzymes, immunoglobulins, antioxidants and antimicrobial peptides (AMPs) in dental plaque, saliva or acquired salivary film).<br>For children up to age 6 years old, the oral cavity of caries-free children (up to 6 years old): 1. Caries-free (CF) children showed lower:<br>a. total protein concentration,<br>b. total antioxidant capacity (TAC),<br>c. serum immunoglobulin-A (S-IgA) concentration<br>d. carbonic anhydrase-VI (CA-VI) activity (for children up to 9 years old),<br>2. caries-affected (CA) children showed lower:<br>a. carbonic anhydrase-VI (CA-VI) concentration (for children up to 9 years old). | NIH Quality Assessment Tool for Observational Cohort and Cross-Sectional Studies (https://www.nhlbi.nih.gov/health-topics/study-quality-assessment-tools) | Yes |
| Ravikumar *et al.*, 2023, Journal of Oral Biology and Craniofacial Research (India) | 1. Cochrane Library<br>2. Embase<br>3. Google Scholar<br>4. Latin America and Caribbean Health Sciences (LILACS)<br>5. MEDLINE<br>6. PubMed<br>7. Scopus<br>8. Manual search<br>9. Grey literature (opengrey.eu)<br>10. Reference lists of included and retrieved articles | Not reported | 4/30/2021 | English only | Cross-sectional studies | Two (2) included studies for meta-analysis were designed as cross-sectional studies. | Are there variations in the physical and chemical properties of saliva in children with and without ECC? | Children: Healthy children less than 6 years of age. | Children: With ECC | Children: Without ECC | Salivary physiochemical properties, inorganic ions, proteins, peptides, and enzymes.<br>1. For children with ECC, as compared to caries free children:<br>a. Salivary IgA values were found to be high and<br>b. Salivary IgG values were found to be high. | Newcastle–Ottawa Scale (NOS) | Yes |

The reviews utilized an array of search restrictions for language which included English and Chinese, English only; and English, German, and Latin-based languages.^21–23^ The primary designs for the three reviews were cross-sectional with respective number of primary studies as seven, fifteen, and two.^21–23^ The primary research question for the three reviews were respectively: 1) “Is there any link between the concentration and/or activity of carbonic anhydrase-VI and the susceptibility to dental caries? ^21^”, 2) “Does the oral cavity of caries-affected individuals has a different protein abundance compared with the oral cavity of caries-free ones? ^22^”, and 3) “Are there variations in the physical and chemical properties of saliva in children with and without ECC?^23^“

The three reviews included the following populations, indicators, comparators, conclusions, assessment tools, and meta-analyses. All three reviews examined populations of healthy children, with two limiting the subject-age to less than nine years old and one to less than age six.^21–23^ The indicator for the three reviews were, respectively, children with “dental caries,” “caries-affected” teeth, and “early childhood caries.^21–23^” The comparator for two reviews were children with “caries-free” teeth and for one review was “without early childhood caries.^21–23^

The conclusions for the three reviews varied with their respective findings by Al-Mahdi *et al*. that “Carbonic anhydrase-VI has lower concentrations and higher activity in children with dental caries;” da Silveira *et al*. that “caries-free children less than nine years old showed lower total protein concentration, total antioxidant capacity (**TAC**), salivary secretory immunoglobulin-A concentration, and carbonic anhydrase-VI activity; and caries-affected children less than nine years old showed lower carbonic anhydrase-VI concentration (for children up to 9 years old);” and Ravikumar *et al*. that, “children with ECC had higher Salivary IgA and Salivary IgG values.^21–23^”

For quality assessment tools, two reviews utilized the Newcastle-Ottawa Scale and one utilized the National Institutes of Health Quality Assessment tool.^21–23^ All three included reviews conducted meta-analyses.

#### Excluded studies

A total of 21 studies that underwent full-text screening were excluded from this umbrella review because they did not fulfill the inclusion and selection criteria.

### Data Analysis and Results of Synthesis

#### Overlap of studies

Table 2 shows the results of the CCA analysis. With three reviews meeting the inclusion criteria, including a total of 22 primary index studies with meta-analyzable data which focused on mean effect differences between caries-affected and caries-free children, the degree of overlap between the reviewed studies was eleven percent, which borders the moderate to high categories.

**Table 2.**
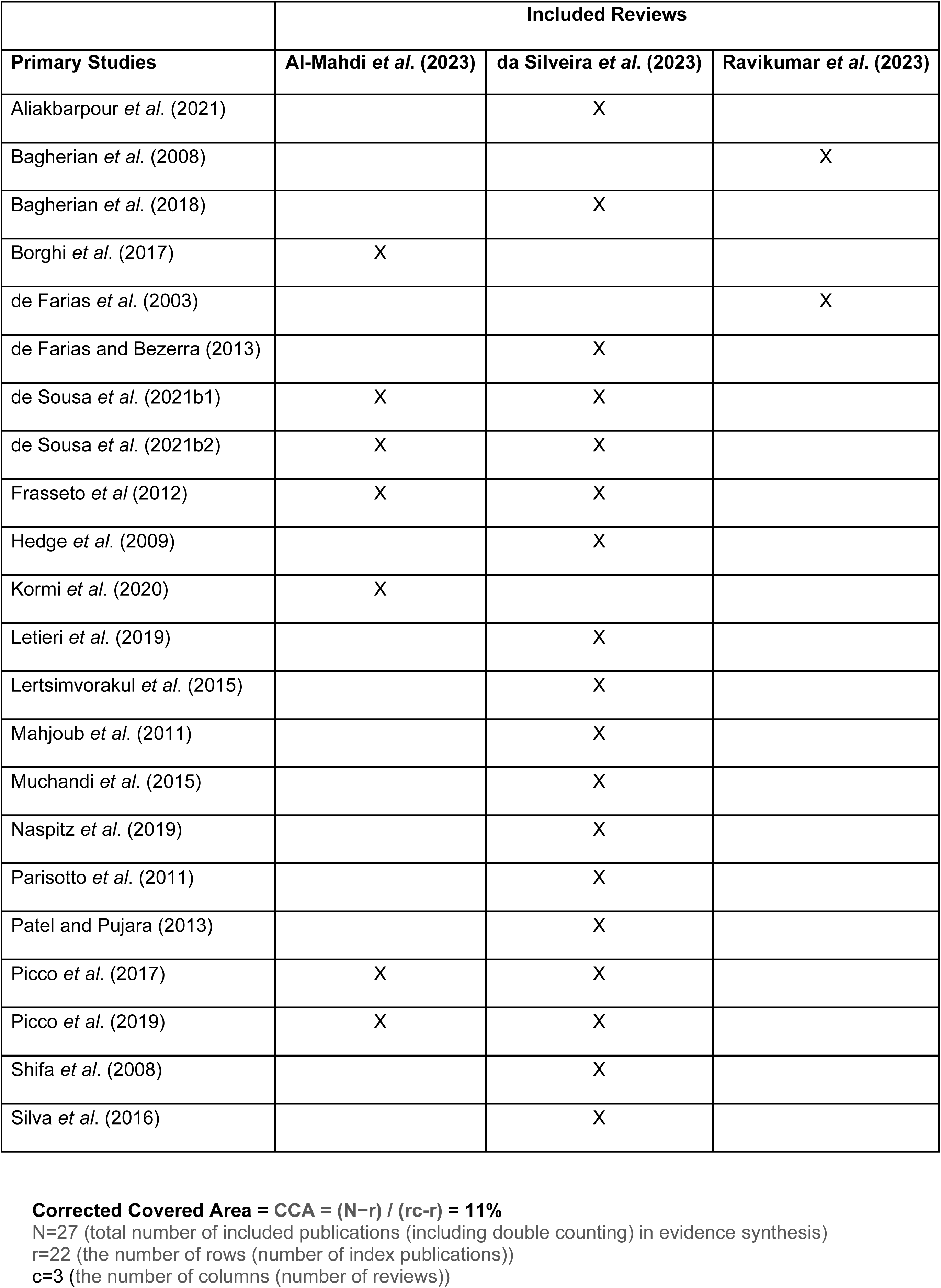
Corrected Covered Area (CCA)

|  | Included Reviews |  |  |
| --- | --- | --- | --- |
| Primary Studies | Al-Mahdi *et al.* (2023) | da Silveira *et al.* (2023) | Ravikumar *et al.* (2023) |
| Aliakbarpour *et al.* (2021) |  | X |  |
| Bagherian *et al.* (2008) |  |  | X |
| Bagherian *et al.* (2018) |  | X |  |
| Borghi *et al.* (2017) | X |  |  |
| de Farias *et al.* (2003) |  |  | X |
| de Farias and Bezerra (2013) |  | X |  |
| de Sousa *et al.* (2021b1) | X | X |  |
| de Sousa *et al.* (2021b2) | X | X |  |
| Frasseto *et al* (2012) | X | X |  |
| Hedge *et al.* (2009) |  | X |  |
| Kormi *et al.* (2020) | X |  |  |
| Letieri *et al.* (2019) |  | X |  |
| Lertsimvorakul *et al.* (2015) |  | X |  |
| Mahjoub *et al.* (2011) |  | X |  |
| Muchandi *et al.* (2015) |  | X |  |
| Naspitz *et al.* (2019) |  | X |  |
| Parisotto *et al.* (2011) |  | X |  |
| Patel and Pujara (2013) |  | X |  |
| Picco *et al.* (2017) | X | X |  |
| Picco *et al.* (2019) | X | X |  |
| Shifa *et al.* (2008) |  | X |  |
| Silva *et al.* (2016) |  | X |  |

**Corrected Covered Area = CCA = (N-r) / (rc-r) = 11%** N=27 (total number of included publications (including double counting) in evidence synthesis) r=22 (the number of rows (number of index publications)) c=3 (the number of columns (number of reviews))

#### Methodological quality of included reviews Quality appraisal

The results of the AMSTAR 2 quality appraisal of the three reviews are presented in Figure 2. Two studies^21,23^ (67 percent) were classified as being of high quality per the AMSTAR 2 rating scale, and one study^22^ (33 percent) was classified as being of low quality.

**Figure 2.**
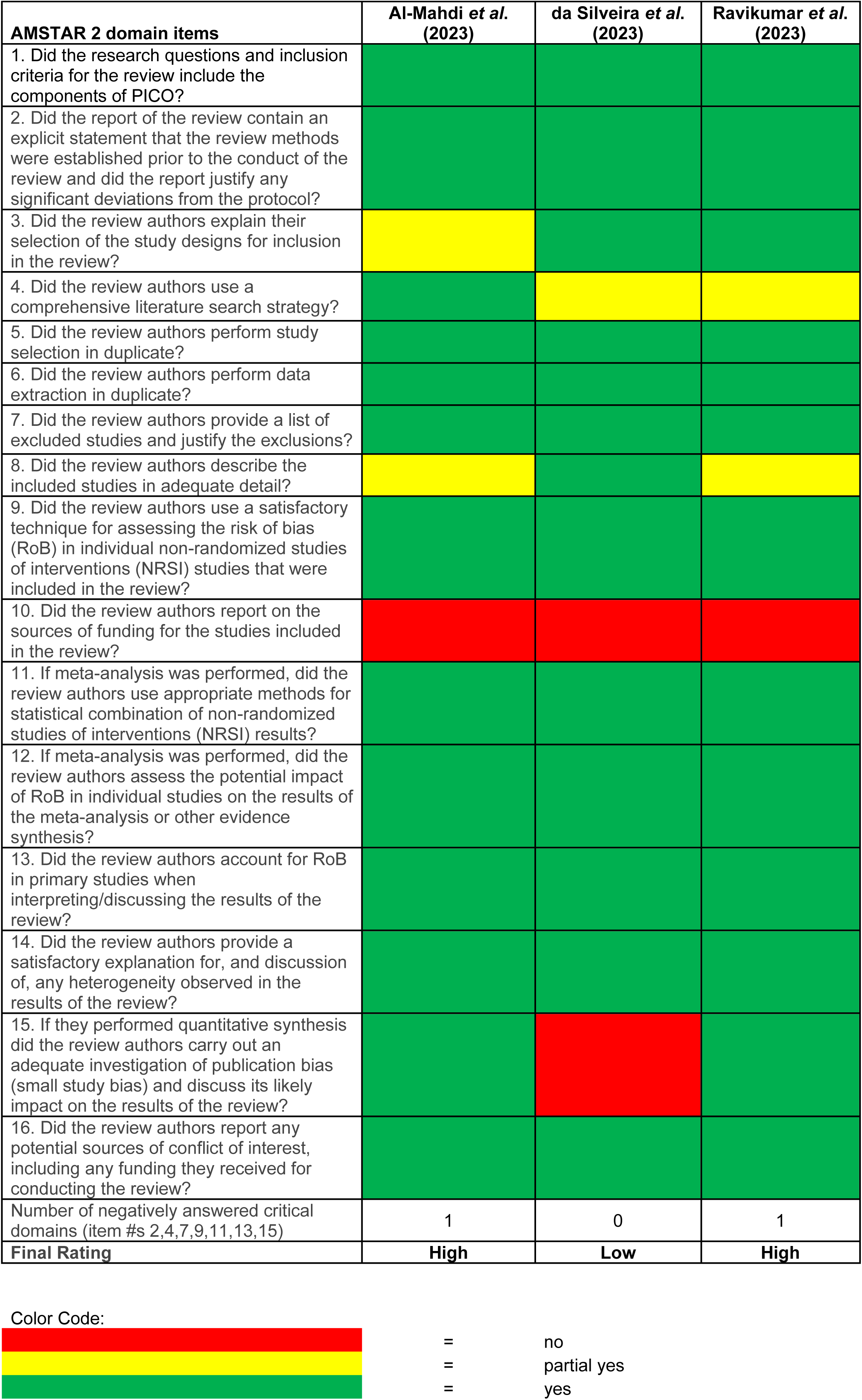
AMSTAR 2 assessment.

One of the reviews^22^ (33 percent) did not conduct the critical investigation of publication bias (item 15), which contributed to the low overall quality assessment. Additionally, the item that was most frequently omitted in the reviews was “item 10,” the non-critical sources of funding for all three (100 percent) of the reviews.^21–23^

#### Risk of Bias (ROB)

AMSTAR 2 was used to assess quality appraisal and the risk of bias. The results are depicted in Figure 2. The results show three (100 percent) of the included reviews had “yes” responses for “item 9” and “item 13” in the AMSTAR 2 assessment. Item 9 asks, “Did the review authors use a satisfactory technique for assessing the RoB in individual studies that were included in the review?” and item 13, “Did the review authors account for RoB in primary studies when interpreting/discussing the results of the review? The risk of bias assessment tools utilized by the one review^22^ included the National Institutes of Health Quality Assessment tool, and two reviews^21,23^ used the Newcastle-Ottawa Scale of which all were found to be satisfactorily addressed.

#### Publication Bias

From the AMSTAR 2 tool (Figure 2), two of the reviews^21,23^ (67 percent) of the included reviews had “yes” responses and one review^22^ (33 percent) had a “no” response for “item 15” in the AMSTAR 2 assessment which asked, *“*If they performed quantitative synthesis did the review authors carry out an adequate investigation of publication bias (small study bias) and discuss its likely impact on the results of the review?” These responses contributed to the final and respective high quality appraisal ratings for the two^21,23^ and low rating for one^22^ included review.

Although the quantitative meta-analysis for both included reviews only totaled less than ten primary, subgroup databases, the funnel-plot analyses showed that the likelihood of publication bias was low for S-IgA concentration (Figure 3.b.), carbonic anhydrase-VI concentration (Figure 3.d.), and carbonic anhydrase-VI activity (Figure 3.f.) because of the symmetry and homogeneity along the center axis with the mean difference nearly equal to zero.^24^

**Figure 3.a.**
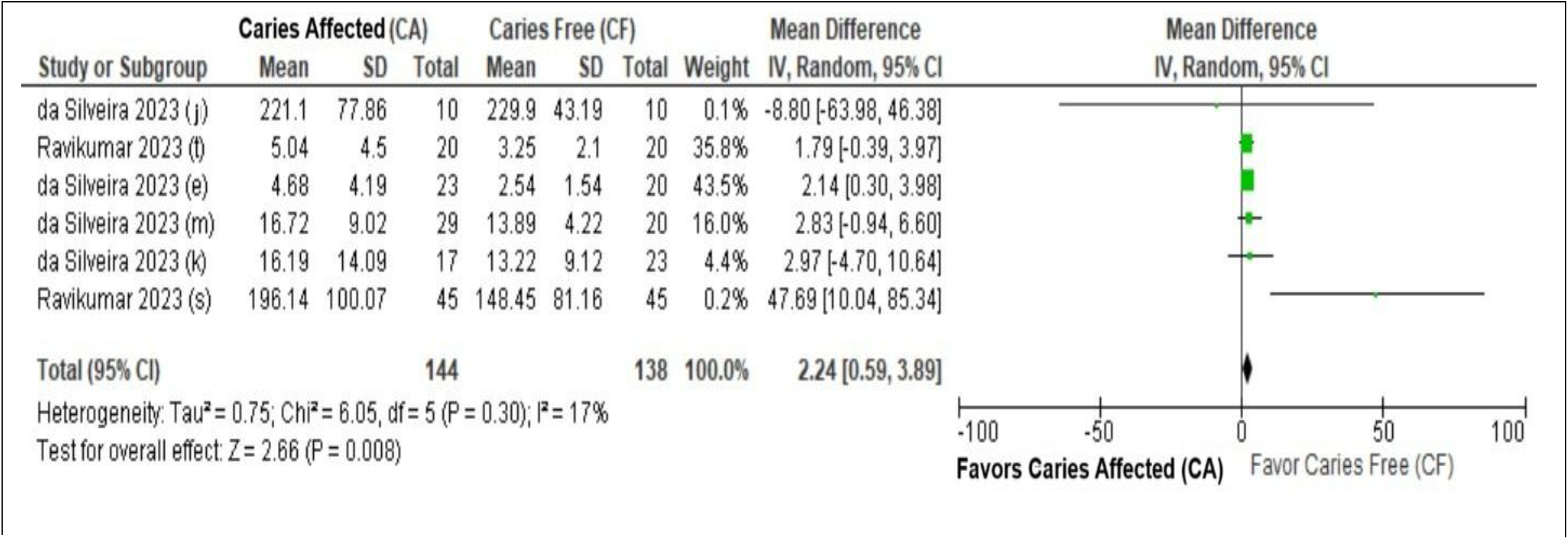
Forest plot for S-IgA Concentration between Caries-Affected (CA) versus Caries-Free (CF) Children.

**Figure 3.b.**
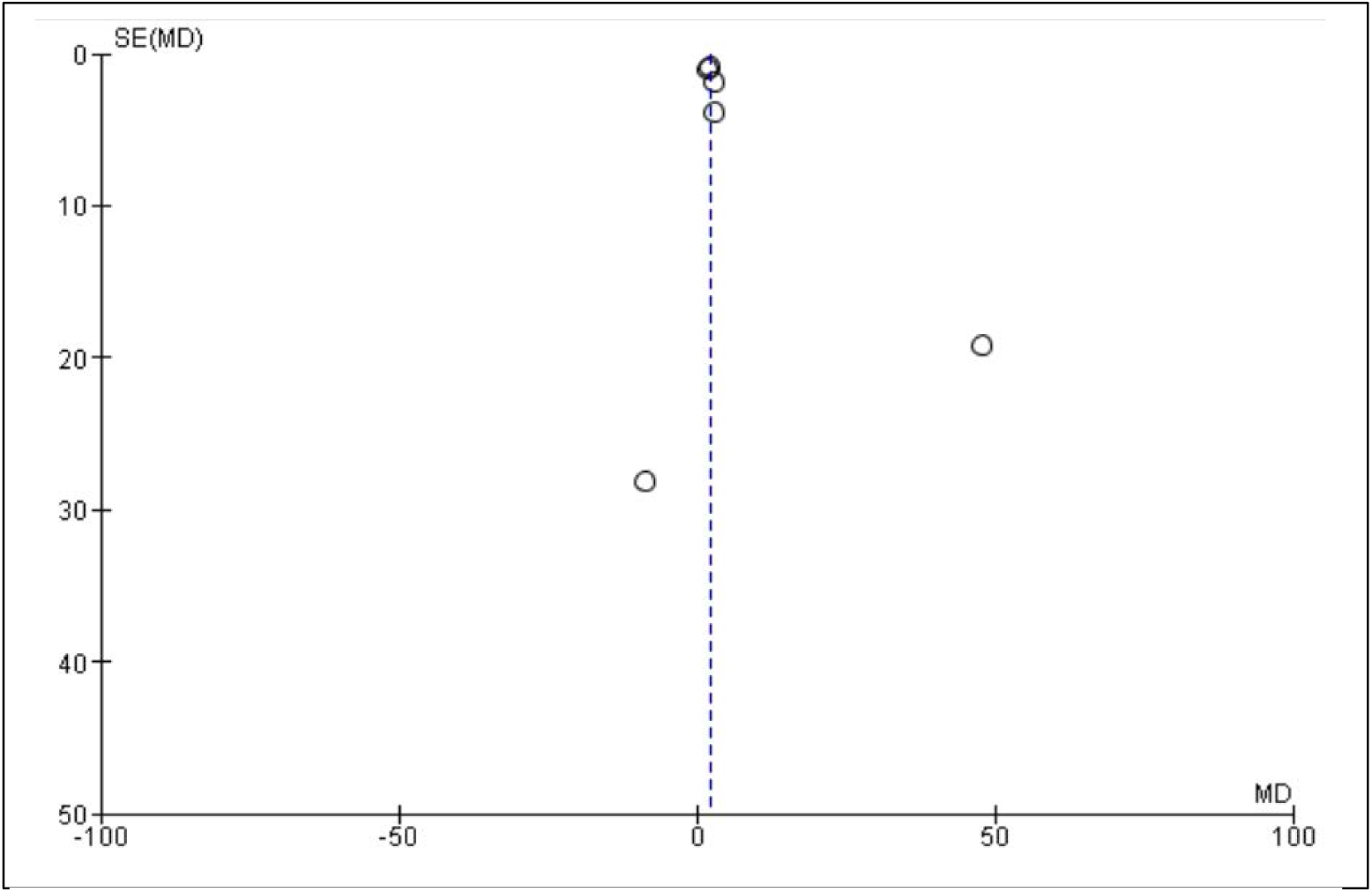
Funnel plot for S-IgA Concentration between Caries-Affected (CA) versus Caries-Free (CF) Children.

**Figure 3.c.**
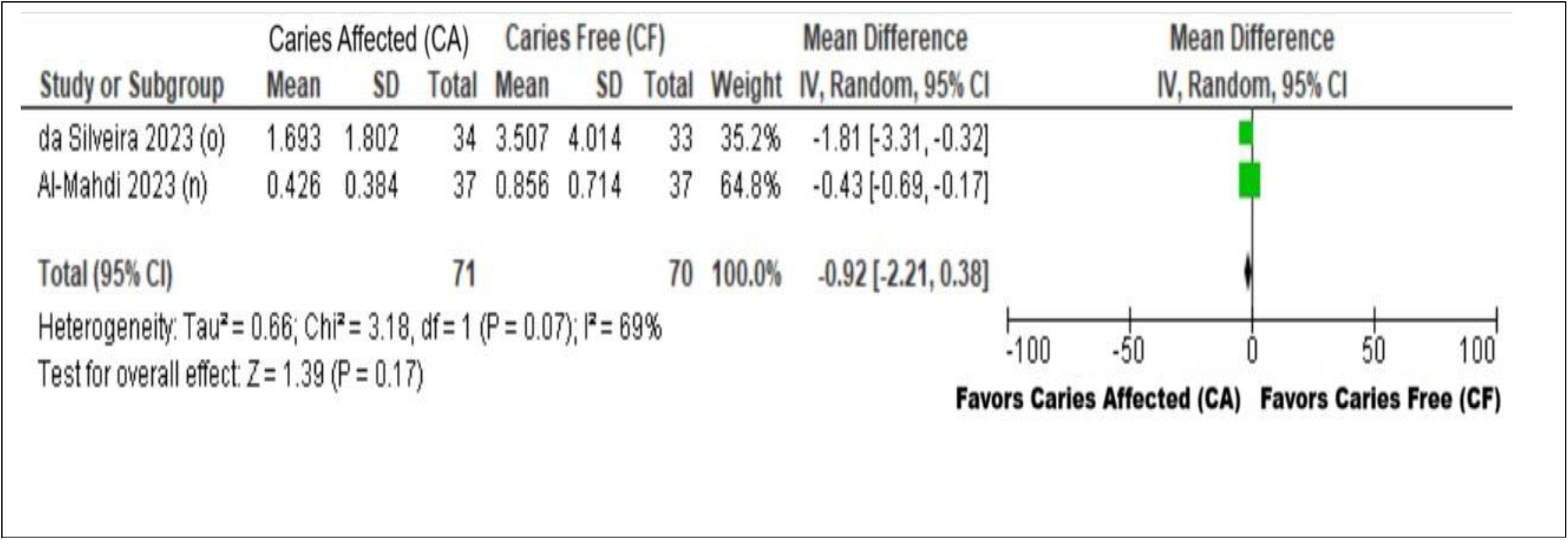
Forest plot for Carbonic anhydrase-VI Concentration between Caries-Affected (CA) versus Caries-Free (CF) Children.

**Figure 3.d.**
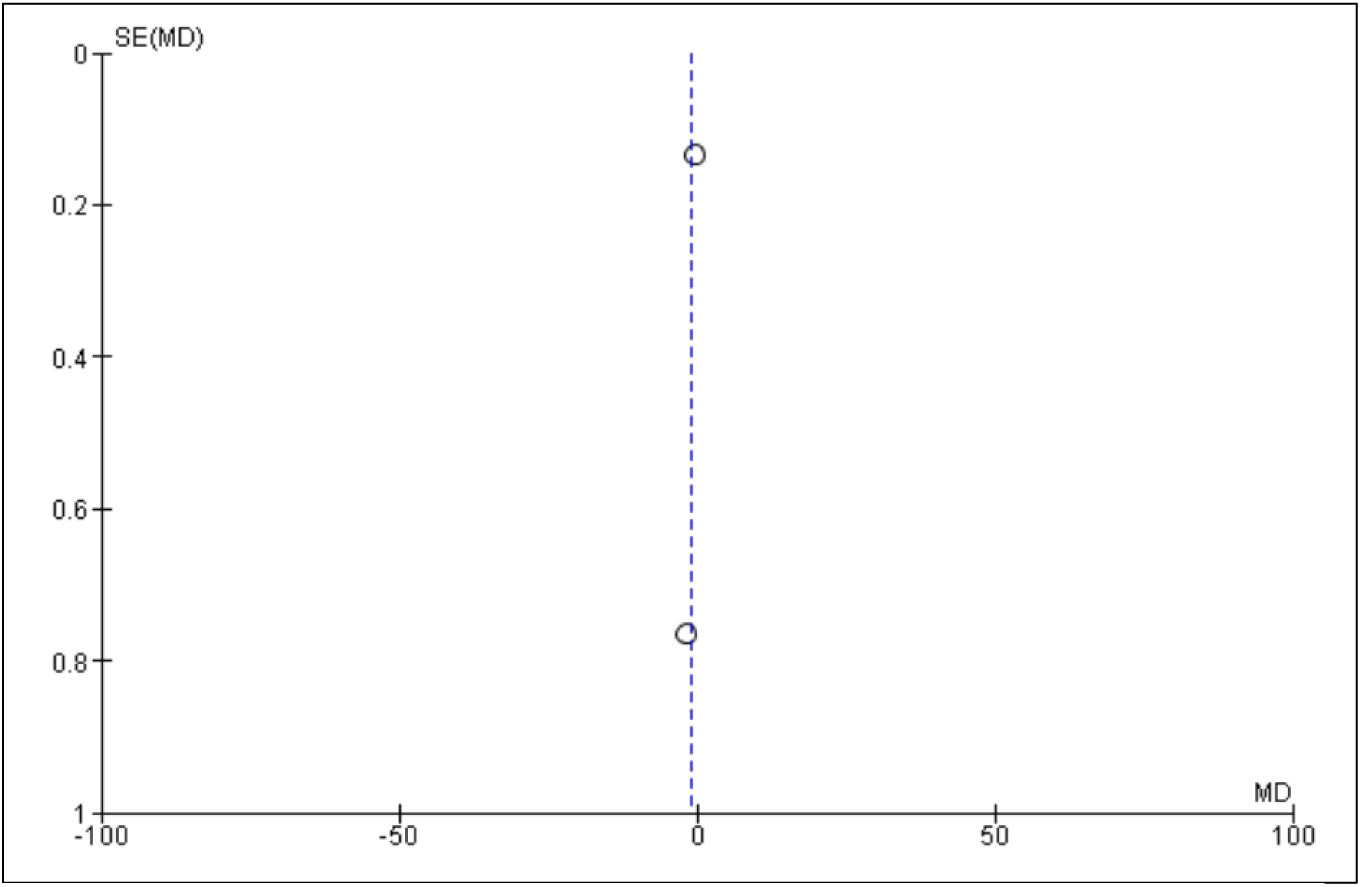
Funnel plot Carbonic anhydrase-VI Concentration between Caries-Affected (CA) versus Caries-Free (CF) Children.

**Figure 3.e.**
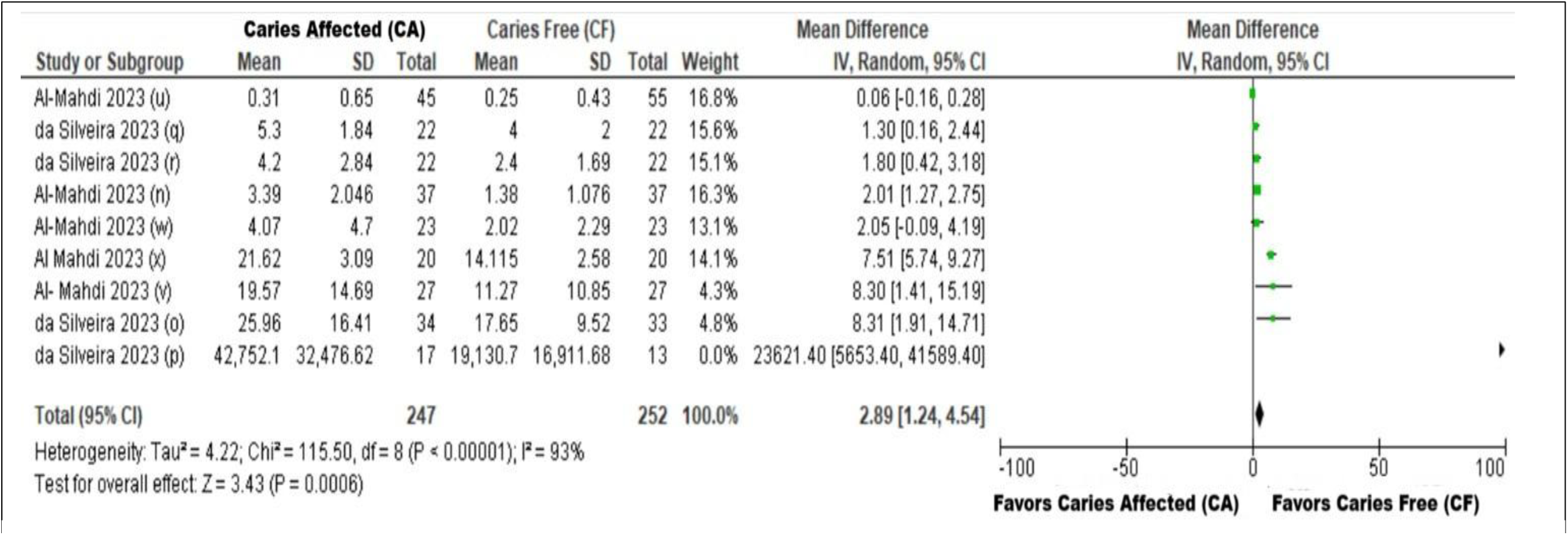
Forest plot for Carbonic Anhydrase Activity between Caries-Affected (CA) versus Caries-Free (CF) in Children.

**Figure 3.f.**
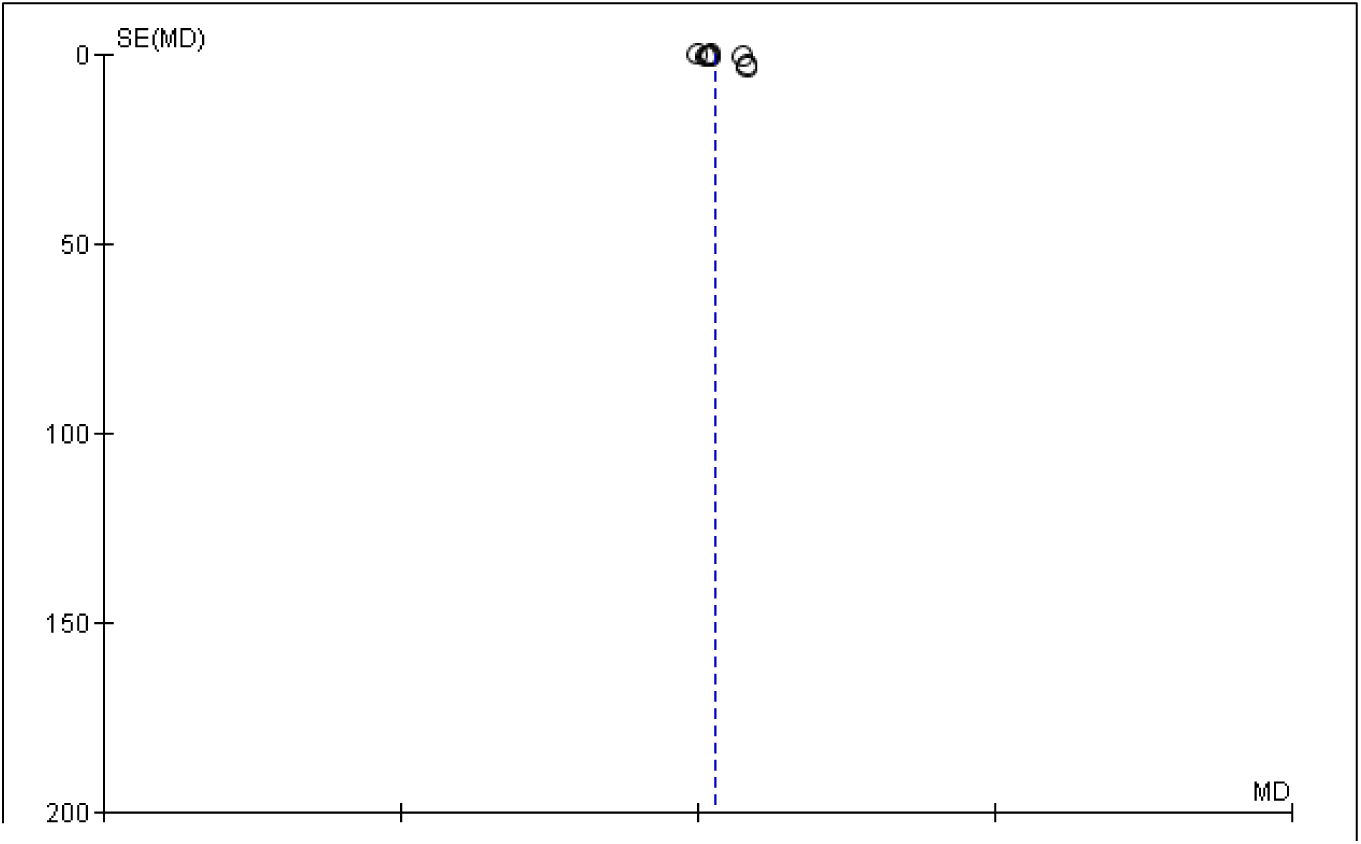
Funnel plot for Carbonic Anhydrase Activity between Caries-Affected (CA) versus Caries-Free (CF) Children.

#### Effects of interventions

##### Meta-analysis summary

Summaries for the meta-analyses of the mean effect of salivary biomarkers between caries-affected and caries-free children are respectively visualized in Figures 3.a., 3.c., and 3.e.

The analysis for S-IgA concentration (Figure 3.a.) included data from a total of six cross-sectional, primary studies for study subgroup analysis, four from da Silveira *et al*. (2023 (e),^25^ (j),^26^ (k),^27^ (m)^28^) data and two from the Ravikumar *et al*. (2023 (s)^29^and (t)^30^) data. In Figure 3.a. the meta-analysis demonstrated an overall pooled effect with the mean difference at 2.24, confidence interval of 0.59 to 3.89, and Z-test for overall effect of 2.66 at P = 0.008. Hence, the higher S-IgA concentration effect favors caries-free children, with statistical significance.

The analysis for carbonic anhydrase-VI concentration (Figure 3.c.) included data from a total of two cross-sectional, primary studies for study subgroup analysis, one from da Silveira *et al*. (2023 (o)^31^), and one from the Al-Mahdi *et al*. (2023 (n)^32^) data. In Figure 3.c. the meta-analysis demonstrated an overall pooled effect with the mean difference at −0.92, confidence interval of 0-2.21 to 0.38, and Z-test for overall effect of 1.39 at P = 0.17. Hence, the higher CA-VI concentration effect favors caries-affected children, without statistical significance.

The analysis for carbonic anhydrase-VI activity (Figure 3.e) included data from a total of nine cross-sectional, primary studies for study subgroup analysis, four from da Silveira *et al*. (2023 (o),^31^ (p),^31^ (q),^33^ (r)^33^) data, and five from the Al-Mahdi *et al*. (2023 (u)^34^, (v)^35^, (w),^35^ (x)^36^) data. In Figure 3.e. the meta-analysis demonstrated an overall pooled effect with the mean difference at 2.89, confidence interval of 1.24 to 4.54, and Z-test for overall effect of 3.43 at P = 0.0006. Hence, the higher carbonic anhydrase-VI activity effect favors caries-free children, with statistical significance.

#### Heterogeneity of Data

Summaries for the meta-analyses of the mean effect of salivary biomarkers between caries-affected and caries-free children are respectively visualized in Figures 3.a., 3.c., and 3.e.

For S-IgA concentration (Figure 3.a.), the test for heterogeneity I² statistic was calculated as 17 percent at P = 0.30. For carbonic anhydrase-VI concentration (Figure 3.c.), the test for heterogeneity I² statistic was calculated as 69 percent at P = 0.07.

For carbonic anhydrase-VI activity (Figure 3.e.), the test for heterogeneity I² statistic was calculated as 93 percent at P < 0.00001. Additionally, from the AMSTAR 2 assessment (Figure 2), all three included reviews had “yes” responses for “item 14” in the AMSTAR 2 assessment, which asked, “Did the review authors provide a satisfactory explanation for, and discussion of, any heterogeneity observed in the results of the review?

Therefore, for S-IgA concentration the heterogeneity is categorized as low and for CA-VI concentration and activity, the heterogeneity is categorized as high.

#### Certainty of Evidence

Table 3 shows the GRADE summary of findings with the overall certainty of evidence ratings and comments for comparing effects of salivary biomarkers between CA and CF children.

**Table 3.** GRADE Summary of Findings.

| Outcomes | Anticipated absolute effects* (95% CI) |  | Relative effect (95% CI) | No of participants (studies) | Certainty of the evidence (GRADE) | Comments |
| --- | --- | --- | --- | --- | --- | --- |
|  | Risk with Caries-Free (CF) Children | Risk with Caries-Affected (CA) Children |  |  |  |  |
| Salivary Secretory Ig-A Concentration (S-IgA) | The mean salivary Secretory Ig-A Concentration was 65.54 | MD 2.24 concentration higher (0.59 higher to 3.89 higher) | - | 282 (6 non-randomized studies) | ⊕⊕⊕○ Moderate<sup>(a)</sup> | Caries-Affected (CA) Children likely increases salivary Secretory Ig-A concentration slightly. |
| Salivary Carbonic Anhydrase Concentration (CA-VI) | The mean salivary Carbonic Anhydrase Concentration was 2.18 | MD 0.92 lower (2.21 lower to 0.38 higher) | - | 141 (2 non-randomized studies) | ⊕⊕⊕○ Moderate<sup>(b)</sup> | Caries-Affected (CA) Children likely reduces salivary Carbonic Anhydrase-VI concentration slightly. |
| Salivary Carbonic Anhydrase Activity (CA-VI) | The mean salivary Carbonic Anhydrase Activity was 3698.30 | MD 2.89 higher (1.24 higher to 4.54 higher) | - | 499 (9 non-randomized studies) | ⊕⊕⊕○ Moderate<sup>(c)</sup> | Caries-Affected (CA) Children likely increases salivary Carbonic Anhydrase-VI activity slightly. |

\*The risk in the intervention group (and its 95% confidence interval) is based on the assumed risk in the comparison group and the relative effect of the intervention (and its 95% CI).
CI: confidence interval; MD: mean difference
GRADE Working Group grades of evidence
High certainty: we are very confident that the true effect lies close to that of the estimate of the effect.
Moderate certainty: we are moderately confident in the effect estimate: the true effect is likely to be close to the estimate of the effect, but there is a possibility that it is substantially different.
Low certainty: our confidence in the effect estimate is limited: the true effect may be substantially different from the estimate of the effect.
Very low certainty: we have very little confidence in the effect estimate: the true effect is likely to be substantially different from the estimate of effect.

For S-IgA concentration, the mean concentration was 65.54, with a mean difference of 2.24 concentration higher (0.59 higher to 3.89 higher) for CA children. The GRADE certainty of evidence was rated as “moderate” confidence with the overarching comment, “Caries-Affected Children likely increases salivary Secretory Ig-A concentration slightly.”

For CA-VI concentration, the mean concentration was 2.18, with a mean difference of 0.92 concentration lower (2.21 lower to 0.38 higher) for CA children. The GRADE certainty of evidence was rated as “moderate” confidence with the overarching comment, “Caries-Affected Children likely reduces salivary Carbonic Anhydrase-VI concentration slightly.”

For CA-VI activity, the mean activity was 3698.30, with a mean difference of 2.89 higher (1.24 higher to 4.54 higher) for CA children. The GRADE certainty of evidence was rated as “moderate” confidence with the overarching comment, “Caries-Affected Children likely increases salivary Carbonic Anhydrase-VI activity slightly.”

## DISCUSSION

### Summary of the main results

#### Key Findings and Relationship to the Hypothesis

The main results from this umbrella review found that with moderate confidence that caries-affected children have slightly higher S-IgA concentration and CA-VI activity, and slightly lower CA-VI concentration in the saliva.

Hence, the findings reject the null hypothesis that there is no difference in the salivary biomarker concentration and/or activity for S-IgA and CA-VI.

### Overall completeness and applicability of evidence

The included reviews in this umbrella review provided information regarding three specific outcomes of interest, S-IgA and CA-VI concentrations and CA-VI activity between CA and CF children. The included reviews incorporated a broad sample of healthy children who were examined in different settings. This, along with findings that were similar among the primary, cross-sectional studies demonstrate the generalizability of the evidence to a broad range of populations.

### Quality of the evidence

Based upon the AMSTAR 2 assessment, meta-analysis findings, and the GRADE assessment, the overall quality of the evidence is presented with moderate confidence.

The AMSTAR 2 assessment showed moderate to high quality ratings for the three included reviews. The meta-analysis findings demonstrated CA children with higher S-IgA and CA-VI activity with statistical significance, and lower CA-VI concentration approaching significance, along with low heterogeneity of the CA-VI data and low risk of publication bias. The GRADE summary of findings provided certainty of evidence with moderate confidence.

### Potential biases in the overview process

This umbrella review adapted the steps according to the Preferred Reporting Items for Systematic Reviews and Meta-Analysis and the Cochrane Handbook for Systematic Reviews of Interventions. Potential bias was mitigated with funnel plot analysis to assess the risk of publication bias and subgroup analysis which showed generally low statistical heterogeneity.

The strengths of this umbrella review include the performance of a systematic and comprehensive search strategy in multiple electronic databases to avoid missing relevant systematic reviews, with meta-analyses, and utilization of two reviewers independently assessed the quality of evidence. The AMSTAR 2 assessment was used as a critical assessment tool for systematic reviews in this umbrella review. Another strength was the extraction of subgroup data from seven primary studies with a large number subjects. Finally, utilization of the GRADE tool with its summary of findings supports the certainty of evidence which was deemed as moderate.

This umbrella review had few limitations which included the moderate to high CCA value where 11 percent of index reviews appeared multiple times across the three included reviews, which may have contributed to the weighting of the results and was mitigated by the utilization of subgroup analysis. Another limitation was that the outcomes were measured as mean difference, instead of relative risk. Despite these limitations, this review provides a comprehensive synthesis of the evidence on the concentration and/or activity of the salivary biomarkers S-IgA and CA-VI between CA and CF children. Dentists who treat children can be more confident that utilizing S-IgA and CA-VI as salivary biomarker screening tests will be likely to benefit and unlikely to cause any harm to children.

### Agreements and disagreements with other studies and/or reviews

#### Salivary S-IgA Concentration in Caries-Affected Children

Salivary S-IgA plays a vital role in oral immunity by preventing bacterial adhesion, neutralizing toxins, and influencing biofilm formation. However, its relationship with dental caries remains inconsistent across studies.

Letieri *et al*. found higher S-IgA levels in caries-affected children, suggesting increased immune activation.^25^ Conversely, Wu *et al*. reported significantly lower S-IgA levels in children with caries, implying a possible protective effect when levels are higher.^37^ Hamid *et al*. examined Down syndrome patients and found inconclusive evidence, likely due to high variability in study designs and caries prevalence in this population.^38^

These discrepancies may arise from variations in saliva collection methods (stimulated vs. unstimulated), age ranges, measurement techniques, and diagnostic criteria for caries. The multifactorial nature of dental caries, including dietary habits, oral hygiene, and microbiome composition, further complicates interpretations.^37^

#### Salivary Carbonic anhydrase-VI Concentration in Caries-Affected Children

Carbonic anhydrase-VI plays a critical role in maintaining oral pH balance and buffering acids that contribute to enamel demineralization.^39^

Picco *et al*. found significantly lower CA-VI concentrations in caries-affected children, suggesting compromised buffering capacity, thus reinforcing the association between lower CA-VI levels and increased caries risk.^31^ However, studies by Szabó *et al.*^40^ and Kivelä *et al*.^41^ noted higher CA-VI concentrations in caries-free children, contrasting with the findings of Picco.^31^

These conflicting results may be due to methodological differences, genetic variations, dietary influences, or saliva flow rates. The role of CA-VI in enhancing saliva’s buffering capacity highlights its significance in caries prevention, though further standardization of measurement techniques is needed.^42^

#### Salivary Carbonic anhydrase-VI Activity in Caries-Affected Children

Beyond concentration, CA-VI enzyme activity affects saliva’s ability to maintain pH homeostasis. Higher enzymatic activity theoretically enhances buffering, but the relationship between CA-VI activity and caries remains unclear.

Frasseto *et al*. and Picco *et al*. reported higher CA-VI activity in children with caries, despite lower total CA-VI concentration.^43,31^ These findings suggest a compensatory increase in response to acidic conditions

This paradox implies that although caries-affected children may experience reduced enzyme levels, the remaining enzyme could exhibit heightened activity to counteract acid production. However, if overall CA-VI concentration remains low, its increased activity may not be sufficient to fully neutralize cariogenic effects.^21^

### Future research

This umbrella review has identified the need to perform more randomized clinical trials in which other outcomes that are important for clinical decision-making are assessed and/or reported. It is also necessary to perform trials in which the number of patients is sufficient enough to detect differences in such outcomes. There is a need for improvement in the design and/or reporting of trials in this area, particularly in those areas related to the randomization method, allocation concealment and blinding.

This umbrella review underscores the need for more rigorous clinical trials with improved methodological consistency. Future studies should focus on validating salivary biomarkers in larger and more diverse populations, standardizing saliva collection and analysis methods, investigating biological mechanisms linking biomarkers to caries progression, exploring multi-biomarker approaches for enhanced diagnostic accuracy, and conducting longitudinal studies to assess biomarker stability over time.

### Implications for Practice

Based on the findings of this current umbrella review, screening children for caries using S-IgA and CA-VI biomarkers could help identify those at risk with minimal harm. While major U.S. commercial medical laboratories provide a wide array of diagnostic services, and based upon the authors’ extensive internet search and telephone communications with the laboratories, it was learned that the laboratories do not yet offer routine assessments for these specific biomarkers.

Dentists should remain updated on the evolving research in salivary diagnostics for ECC as the future integration of saliva-based diagnostic tools into dental practice could facilitate early intervention and personalized preventive strategies.^37^

### Implications for Dental Education and Policy

The potential of salivary biomarkers in pediatric dentistry carries important implications for education and policy. Dental curricula should incorporate training on salivary biochemistry, emphasizing the diagnostic relevance for S-IgA and CA-VI.^37^ Professional organizations should also promote research initiatives focused on biomarker validation and point-of-care screening tools.

Once standardized, salivary biomarkers could be incorporated into national oral health programs to improve early caries detection and risk assessment.^2^ Public health agencies should explore integrating saliva-based diagnostics into preventive care models, while policymakers should prioritize research funding for non-invasive screening technologies.^37^

## CONCLUSIONS

Based upon the results of this study, the following conclusions can be made:

1. Caries-affected children under age nine years old exhibit higher S-IgA concentration and CA-VI activity, and lower CA-VI concentration in their saliva.
2. Dentists who treat children can be more confident that utilizing S-IgA and CA-VI as salivary biomarker screening tests will be likely to benefit and unlikely to cause any harm to children.
3. Until U.S. commercial medical laboratories provide standard diagnostic services with saliva-based tests such as specific measurements of S-IgA concentration, carbonic anhydrase-VI concentration, and carbonic anhydrase-VI activity, dentists will need to wait before non-invasive testing with salivary biomarkers is available to identify children at risk for caries-affected dentition.

## Data Availability

All data produced in the present work are contained in the manuscript

## ACKNOWLEDGMENTS

The authors extend appreciation and acknowledgment for funding to the Hansjorg Wyss Department of Plastic Surgery, NYU Langone Hospitals, New York. This research was facilitated by Gemini Deep Research artificial intelligence (**AI**) tool (Google, Inc., Mountain View, CA), accessed on May 11, 2025, at the link, https://gemini.google.com/app. The authors acknowledge the tool’s assistance for the introduction section to identify key themes and for the discussion section to supplement the findings of other studies, which support or contradict the results, and enhance the implications of this umbrella review. The authors affirm that the original intent and meaning of the content remain unaltered during editing and that the AI tools had no involvement in shaping the intellectual content of this work. The authors assume full responsibility for upholding the integrity of the content presented in this manuscript.

## REFERENCES

1. Surwaich, A., Maqbool, A., Majeedaro, S. A., Ali, A. T., Arain, B., Anwar, K., Ahsan, S., & Ejaz, M. (2024). Evaluating Preventive Health Strategies: Salivary Biomarkers as Non-Invasive Indicators of Caries Risk in School Children: Salivary Biomarkers for Caries Risk in Children. Pakistan Journal of Health Sciences, 5(10), 248–257. 10.54393/pjhs.v5i10.2012

2. American Academy of Pediatric Dentistry. Policy on early childhood caries (ECC): Consequences and preventive strategies. The Reference Manual of Pediatric Dentistry. Chicago, Ill.: American Academy of Pediatric Dentistry; 2024:89–92.

3. Thanh MTG, Van Toan N, Toan DTT, Thang NP, Dong NQ, Dung NT, Hang PTT, Anh LQ, Tra NT, Ngoc VTN. Diagnostic Value of Fluorescence Methods, Visual Inspection and Photographic Visual Examination in Initial Caries Lesion: A Systematic Review and Meta-Analysis. Dentistry Journal. 2021; 9(3):30. 10.3390/dj9030030

4. Nahas M, Sfeir E. Salivary Immunoglobulin A and *Streptococcus mutans* Levels among Lebanese Preschool Children with Early Childhood Caries. J Contemp Dent Pract. 2020;21(9):1012–1017. Published 2020 Sep 1

5. Ahmad P, Hussain A, Carrasco-Labra A, Siqueira WL. Salivary Proteins as Dental Caries Biomarkers: A Systematic Review. Caries Res. 2022;56(4):385–398. doi:10.1159/000526942

6. Albagieh H, Alshehri AZ, Alduraywishi AS, et al. Evaluation of Salivary Diagnostics: Applications, Benefits, Challenges, and Future Prospects in Dental and Systemic Disease Detection. Cureus. 2025;17(1):e77520. Published 2025 Jan 16. doi:10.7759/cureus.77520

7. Hu CC, Wang SG, Gao Z, et al. Emerging salivary biomarkers for early detection of oral squamous cell carcinoma. World J Clin Oncol. 2025;16(4):103803. doi:10.5306/wjco.v16.i4.103803

8. Constantin V, Luchian I, Goriuc A, Budala DG, Bida FC, Cojocaru C, Butnaru O-M, Virvescu DI. Salivary Biomarkers Identification: Advances in Standard and Emerging Technologies. Oral. 2025; 5(2):26. 10.3390/oral5020026

9. Zhang Y, Huang S, Jia S, et al. The predictive power of saliva electrolytes exceeds that of saliva microbiomes in diagnosing early childhood caries. J Oral Microbiol. 2021;13(1):1921486. Published 2021 May 13. doi:10.1080/20002297.2021.1921486

10. Zafar M, Levy SM, Warren JJ, Xie XJ, Kolker J, Pendleton C. Prevalence of non-cavitated lesions and progression, regression, and no change from age 9 to 23 years. J Public Health Dent. 2022;82(3):313–320. doi:10.1111/jphd.12538

11. Hultquist AI, Brudin L, Bågesund M. Early childhood caries risk assessment in 1-year-olds evaluated at 6-years of age. Acta Odontol Scand. 2021;79(2):103–111. doi:10.1080/00016357.2020.1795247

12. Stewart L, Moher D, Shekelle P. Why prospective registration of systematic reviews makes sense. Syst Rev. 2012;1:7. Published 2012 Feb 9. doi:10.1186/2046-4053-1-7

13. Page MJ, McKenzie JE, Bossuyt PM, et al. The PRISMA 2020 statement: an updated guideline for reporting systematic reviews. BMJ. 2021;372:n71. Published 2021 Mar 29. doi:10.1136/bmj.n71

14. Pollock M, Fernandes RM, Becker LA, Pieper D, Hartling L. Chapter V: Overviews of Reviews [last updated August 2023]. In: Higgins JPT, Thomas J, Chandler J, Cumpston M, Li T, Page MJ, Welch VA (editors). Cochrane Handbook for Systematic Reviews of Interventions version 6.5. Cochrane, 2024. Available from www.training.cochrane.org/handbook.

15. Pitts NB, Ekstrand KR; ICDAS Foundation. International Caries Detection and Assessment System (ICDAS) and its International Caries Classification and Management System (ICCMS) - methods for staging of the caries process and enabling dentists to manage caries. Community Dent Oral Epidemiol. 2013;41(1):e41–e52. doi:10.1111/cdoe.12025

16. Klein H, Palmer CE, Knutson JW. Studies on dental caries. Public Health Rep. 1938;53:751–765. https://mmclibrary.com/wp-content/uploads/2021/12/Sex-Differences-in-Dental-Caries-Experience-of-Elementary-School-Children.pdf. Accessed on May 13, 2025.

17. Pieper D, Antoine SL, Mathes T, Neugebauer EA, Eikermann M. Systematic review finds overlapping reviews were not mentioned in every other overview. J Clin Epidemiol. 2014;67(4):368–375. doi:10.1016/j.jclinepi.2013.11.007

18. Shea BJ, Reeves BC, Wells G, et al. AMSTAR 2: a critical appraisal tool for systematic reviews that include randomised or non-randomised studies of healthcare interventions, or both. BMJ. 2017;358:j4008. Published 2017 Sep 21. doi:10.1136/bmj.j4008

19. Higgins JP, Thompson SG, Deeks JJ, Altman DG. Measuring inconsistency in meta-analyses. BMJ. 2003;327(7414):557–560. doi:10.1136/bmj.327.7414.557

20. Bezerra CT, Grande AJ, Galvão VK, Santos DHMD, Atallah ÁN, Silva V. Assessment of the strength of recommendation and quality of evidence: GRADE checklist. A descriptive study. Sao Paulo Med J. 2022;140(6):829–836. doi:10.1590/1516-3180.2022.0043.R1.07042022

21. Al-Mahdi R, Al-Sharani H, Al-Haroni M, Halboub E. Associations of the activity and concentration of carbonic anhydrase VI with susceptibility to dental caries: A systematic review and meta-analysis. Clin Exp Dent Res. 2023;9(2):358–367. doi:10.1002/cre2.723

22. da Silveira EG, Prato LS, Pilati SFM, Arthur RA. Comparison of oral cavity protein abundance among caries-free and caries-affected individuals-a systematic review and meta-analysis. Front Oral Health. 2023;4:1265817. Published 2023 Sep 15. doi:10.3389/froh.2023.1265817

23. Ravikumar D, Ramani P, Gayathri R, Hemashree K, Prabhakaran P. Physical and chemical properties of saliva and its role in Early Childhood caries - A systematic review and meta-analysis. J Oral Biol Craniofac Res. 2023;13(5):527–538. doi:10.1016/j.jobcr.2023.05.011

24. Sterne JA, Sutton AJ, Ioannidis JP, et al. Recommendations for examining and interpreting funnel plot asymmetry in meta-analyses of randomised controlled trials. BMJ. 2011;343:d4002. Published 2011 Jul 22. doi:10.1136/bmj.d4002

25. Letieri ADS, Freitas-Fernandes LB, Valente APC, Fidalgo TKDS, de Souza IPR. Longitudinal evaluation of salivary IgA-S in children with early childhood caries before and after restorative treatment. J Clin Pediatr Dent. (2019) 43(4):239–43. 10.17796/1053-4625-43.4.3

26. Shifa S, Muthu MS, Amarlal D, Rathna Prabhu V. Quantitative assessment of IgA levels in the unstimulated whole saliva of caries-free and caries-active children. J Indian Soc Pedod Prev Dent. (2008) 26(4):158–61. 10.4103/0970-4388.44031

27. Parisotto TM, King WF, Duque C, Mattos-Graner RO, Steiner-Oliveira C, Nobre-Dos-Santos M, et al. Immunological and microbiologic changes during caries development in young children. Caries Res. (2011) 45(4):377–85. 10.1159/000330230

28. Naspitz GM, Nagao AT, Mayer MP, Carneiro-Sampaio MM. Anti-Streptococcus mutans antibodies in saliva of children with different degrees of dental caries. Pediatr Allergy Immunol. (1999) 10(2):143–8. 10.1034/j.1399-3038.1999.00026.x

29. Bagherian A., Jafarzadeh A., Rezaeian M., Ahmadi S., Rezayati M. Comparison of the salivary immunoglobulin concentration levels between children with early childhood caries and caries-free children. Iran J Immunol. 2008;5:217–221

30. De Farias D.G., Bezerra A.C. Salivary antibodies, amylase and protein from children with early childhood caries. Clin Oral Invest. 2003;7:154–157. doi: 10.1007/s00784-003-0222-7

31. Picco, D.C.R., Lopes, L.M., Steiner-Olivera, C., & Nobre Dos Santos, M. (2022). The protective potential of carbonic anhydrase VI (CA VI) against tooth decay in children: A systematic review of the literature. Journal of Clinical Advances in Dentistry, 6, 021–027.

32. Picco, D.C.R., Lopes, L.M., Rocha Marques, M., Line, S.R.P., Parisotto, T.M., & Nobre Dos Santos, M. (2017). Children with a higher activity of carbonic anhydrase VI in saliva are more likely to develop dental caries. Caries Research, 51(4), 394–401. 10.1159/000470849

33. Sousa ET, Lima-Holanda AT, Nobre-Dos-Santos M. Carbonic anhydrase VI activity in saliva and biofilm can predict early childhood caries: a preliminary study. Int J Paediatr Dent. (2020) 31(3):361–71. 10.1111/ipd.12717

34. Borghi, G. N., Rodrigues, L. P., Lopes, L. M., Parisotto, T. M., Steiner-Oliveira, C., & Nobre-Dos-Santos, M. (2017). Relationship amongαamylase and carbonic anhydrase VI in saliva, visible biofilm, and early childhood caries: A longitudinal study. International Journal of Paediatric Dentistry, 27(3), 174–182. 10.1111/ipd.12249

35. de Sousa, E.T., Lima-Holanda, A.T., Sales, L.S., & Nobre-Dos-Santos, M. (2021b). Combined effect of starch and sucrose on carbonic anhydrase VI activity in saliva and biofilm of children with early childhood caries. Exposure to starch and sucrose alters carbonic anhydrase VI activity in saliva and biofilm. Clinical Oral Investigations, 25(5), 2555–2568. 10.1007/s00784-020-03567-z

36. Kormi, E., Pani, S., AlNatsha, J., & Stankeviciene, R. (2020). Association between salivary carbonic anhydrase 6 levels and early childhood caries in children with chronic bronchial asthma [Unpublished Data].

37. Wu Z, Gong Y, Wang C, Lin J, Zhao J. Association between salivary s-IgA concentration and dental caries: A systematic review and meta-analysis. Biosci Rep. Published online December 8, 2020. doi:10.1042/BSR20203208

38. Hamid H, Adanir N, Asiri FYI, Abid K, Zafar MS, Khurshid Z. Salivary IgA as a Useful Biomarker for Dental Caries in Down’s Syndrome Patients: A Systematic Review and Meta-analysis. Eur J Dent. 2020;14(4):665–671. doi:10.1055/s-0040-1716443

39. Antonelli R, Massei V, Ferrari E, et al. Salivary Diagnosis of Dental Caries: A Systematic Review. Curr Issues Mol Biol. 2024;46(5):4234–4250. Published 2024 May 2. doi:10.3390/cimb46050258

40. Szabó I. Carbonic anhydrase activity in the saliva of children and its relation to caries activity. Caries Res. 1974;8(2):187–191. doi:10.1159/000260107

41. Kivelä J, Parkkila S, Parkkila AK, Rajaniemi H. A low concentration of carbonic anhydrase isoenzyme VI in whole saliva is associated with caries prevalence. Caries Res. 1999;33(3):178–184. doi:10.1159/000016514

42. Makawi Y, El-Masry E, El-Din HM. Salivary carbonic anhydrase, pH and phosphate buffer concentrations as potential biomarkers of caries risk in children. J Unexplored Med Data 2017;2:9–15. Accessed May 17, 2025. https://www.oaepublish.com/articles/2572-8180.2016.07.

43. Frasseto F, Parisotto TM, Peres RC, Marques MR, Line SR, Nobre Dos Santos M. Relationship among salivary carbonic anhydrase VI activity and flow rate, biofilm pH and caries in primary dentition. Caries Res. 2012;46(3):194–200. doi:10.1159/000337275

